# How to Monitor Physical activity in pregnant women? Questionnaire and accelerometer: stages of building a virtual assistant

**DOI:** 10.64898/2026.05.07.26343713

**Authors:** Gleici da Silva Castro Perdona, Carla Micheli da Silva, Tiago Chiaveri da Costa, Rafael Fazio, Nathan Zanutto, Carlos Capellini, Leonardo Facci

## Abstract

**Introduction:** Physical activity during pregnancy can be tracked directly by accelerometer measurements and indirectly by validated questionnaires. Considering the advancement of the Internet of Things (IOT), managing and/or monitoring physical activities can be better explored to analyze individuals, as well as indirectly compare the intensity and domains of physical activities carried out by pregnant women. The project, called “EVA”(Expert Virtual Assistant), suggests combining several fields of knowledge to obtain better information about physical activity during pregnancy, surpassing the claim made in previous research that studying and measuring the duration of daily physical activities in pregnant women is a challenge.

**Objective:** In the present study, we present the results of the first stage of the EVA project, which aims to develop a Virtual Assistant (VA) in Portuguese, providing examples of health management features for monitoring Physical Activity measurements for pregnant women assisted in the Unified Health System (SUS) and the adaptation of the Pregnancy Physical Activity Questionnaire (PPAQ).

**Methods and Analysis:** The methods used were developed in two stages: adapting the physical activity questionnaire and building the Virtual Assistent to monitor physical activities. Thirty pregnant women who used the Unified Health System (SUS) in the city of Ribeirão Preto, Brazil participated in the study. The pregnant women wore sensor wristbands (accelerometers) and answered the sociodemographic, lifestyle and physical activity questionnaires via an application developed for this study.

**Results:** The questionnaire used was the PPAQ adapted for Brazilian pregnant women. The most important changes were in the occupational domain for the house cleaning and in sedentary behavior activities. In the pilot study, it was observed that pregnant women spend more energy at home and in light and moderate intensity activities. textbfConclusion:This study made important contributions to evaluating PA in pregnant women. The proposal and studies for the construction of the AV-EVA, the inclusion of a specific occupational domain for pregnant women with domestic occupations and the new cutoff points for PA intensity measurements obtained via accelerometers.

## INTRODUCTION

The potential of the Internet of Things (IoT) for individual health management can also be explored for pregnancy care. The IoT is a network of devices that are embedded with sensors and network connections that can collect and transmit data. IoT implications include the fact that it is inexpensive and there are innovative high-quality wearable or remote biosensors; and it generates a large amount of data in real time, for example, related to the health of thousands of consumers (De Paula, 2014) In this context, developing a Virtual Assistant (VA) in Portuguese providing examples of health management features for monitoring Physical Activity measurements for pregnant women gave rise to the Expert Virtual Assistant nicknamed EVA project, which added to its development physical activity monitoring, such as adapting a questionnaire to measure the intensity and domains of physical activities carried out by pregnant women. Physical activity (PA) is important for full human development and should be practiced at all stages of life and at different times, such as when moving from one place to another, during work or when doing household chores.

During pregnancy, regular PA can provide several health benefits for the mother and fetus, such as reduced depression symptoms and lower risk of excessive gestational weight gain, it reduces the risk of developing gestational diabetes mellitus (Silva et al., 2021; Han et al., 2020; ACOG, 2020), preeclampsia and hypertension, which is the main cause of maternal mortality in Brazil (12 to 22% of pregnant women)(Santini et al., 2017; Santos et al., 2020).

The American College of Obstetricians and Gynecologists (ACOG) recommends that pregnant women should do at least 20-30 min a day, or 150 min per week, of moderate intensity aerobic activity every day of the week, or nearly every day in the absence of obstetric or medical complications or contraindications (ACOG, 2020). To verify the PA level of pregnant women, questionnaires are the most widely used instruments in epidemiological studies due to ease of application and low cost (Silva et al., 2021; Sattler et al., 2018; Younis et al., 2021). It is essential to use specific questionnaires with pregnant women, which are of good quality and that accurately measure the PA in this population as questionnaires designed for the general population do not include household chores and other activities that are commonly done by pregnant women(Santini et al., 2017; Han et al., 2020). The Preqnancy Physical Activity Questionnaire (PPAQ) was developed and validated by Chasan-Taber et al. (2004) that measures PA during pregnancy and includes questions that encompass all PA domains (household, transportation, occupational, leisure) showing good quality and strong evidence to measure PA in pregnant women.

The PPAQ has been translated and adapted for various populations, such as Korean (Han et al., 2020), Arabic (Papazian et al., 2020), Polish (Krzepota et al., 2017) and Brazilian (Silva et al., 2015). However, even considering adapting the PPAQ to the Brazilian population, it is important to consider the specific characteristics of the sample to be studied when asking participants to respond to a questionnaire to assess PA. A particular semi-structured questionnaire was applied and validated in 34 pregnant women in the study conducted by Takito et al. (2008) in Brazil. The questionnaire was developed based on the Leisure Time Physical Activity (LPTA) and Household Activities and International Physical Activity Questionnaire (IPAQ) long version, as well as Wildschut et al. (1993), Taylor et al. (1978), and Tavares et al. (2009).

For all the authors, being part of the population of pregnant women is a specific characteristic, as pregnant women are called, in terms of PA, as a sedentary population.

Questioning this hypothesis, we propose a study that considers evaluating both, a PA by questionnaire and PA by an accelerometer, comparing the results and determining the cutoff points to classify the PA intensity. Thus, this study aims to present the results of the PPAQ adaptation and the pilot study carried out in the field using the VA, called EVA. More details about this project in Portuguese can be found at: www.eva.fmrp.usp.br.

## METHODS

The project EVA was divided into the following stages: The first stage (STAGE I) consisted of defining the questionnaire; Focus group, Pilot study and building the Virtual Assistant (VA). The second stage (STAGE II) consisted of validating the instrument of the questionnaire adapted by this study with the accelerometer. In this paper, we will present the results of the first stages of the EVA project (STAGE I). To contextualize we present the stages of the project.

### STAGE I: Adapting the Pregnancy Physical Activity Questionnaire

This part of the study consisted of four phases: **Phase I** - Defining the instrument for the study, **Phase II** - Focus group and **Phase III** - building the VA and **Phase IV** Pilot study.

#### Phase I: Defining the questionnaire

According to Sattler et al. (2018), when choosing a questionnaire for pregnant women, some properties of important measures should be considered, such as: (1) the questionnaire should cover all PA domains (household, occupational, leisure, transportation); (2) it should measure the duration and frequency of PA, (3)the recall period should be the last week (or last seven days) in the current trimester, due to the lack of the specific Metabolic Equivalent Task (MET) intensities for pregnant women and the changes in energy expenditure during pregnancy, (4) the use of total time is recommended when assessing total PA. Considering the results of the study by (Sattler et al., 2018), the PPAQ proved to have sufficient measurement properties to measure PA in various populations (Han et al., 2020; Krzepota et al., 2017; Papazian et al., 2020; Sattler et al., 2018; Silva et al., 2015). In Brazil, the study by (Silva et al., 2015) translated the PPAQ into “Brazilian Portuguese”, which they called the Physical Activity Questionnaire for Pregnant Women (PPAQW). The PPAQ, which was originally written in English, consists of 36 questions in total and 32 of these questions address PA and demonstrated sufficient validity and reliability (total activities, ICC= 0.78, sedentary activity, ICC= 0.79, moderate activity, ICC= 0.82). (Chasan-Taber et al., 2004) The PAQPW (Silva et al., 2015) consists of 33 questions, two of which address information about the woman’s last menstrual period and the remaining 31 refer to PA in all domains. One question from the PPAQW was withdrawn as it was completely out of the context of Brazilian pregnant women. However, there is no study concerning validation with a direct instrument Da Silva and Perdona (2026).

Considering the socioeconomic and cultural diversity of the Brazilian population due to the large territorial extension, it was considered necessary to verify it using a qualitative study (focus group) if there was a need for an adaptation of the translated questionnaire, we will call: B-PPAQ (The full version of the B-PPAQ questionnaire in Portuguese is available upon request from the corresponding author).

#### Phase II: Focus Group

The focus group collects information based on communication and interaction between its participants Eeg-Olofsson et al. (2020). The main objective is to collect detailed information from a group of selected individuals concerning their characteristics and representativeness, providing an open and accessible debate around a topic of common interest Chopra et al. (2021). The focus group met in a Health Facility (HF), which at the time was offering a pre-natal course for pregnant women who used the Unified Health System (SUS) in the city of Ribeirão Preto.

The technical team consisted of a psychologist, a physical educator and the coordination researcher of the project. The sessions, lasting 40 minutes, held at the end of the pre-natal course, consisted of 4 meetings. Three meetings were with the pregnant women and a follow up with specialists to discuss impressions and necessary adaptations. Pregnant women over 18 years of age and of different gestational weeks participated in the focus group to ensure heterogeneity in the sample according to the study population. The pregnant women were invited to participate in the focus group intentionally, free from any coercion. An informed consent form was given to the Basic Health Facility coordination team and the pregnant women. The wording and order of the questions discussed in the focus group were based on the PPAQ and the PPAQW. Therefore, in the first meeting, the household domain was addressed; in the second meeting, the occupational domain; and in the third meeting, the leisure and transportation domains. Due attention was given to ensuring that the questions were in plain language, without ambiguities and expressed in the first person. All procedures of the research were clarified and participation was voluntary, upon signing the Term of Consent. Data collection tool place on August 27,2019 and November 29,2019.

#### Phase III: Building the Virtual Assistant

The Virtual Assistant started being built parallel to the focus group. VA was developed in Python3 language using a database structure (MySQL); an application is used that connected the information from the proposed questionnaire in electronic form; it had a connection with a pregnancy recording device and a user-computer interface with voice command. For integration and data acquisition via IoT, accelerometers specialized in PA, which captured the relevant measures, PA intensity, through variations of the data generated in the triaxial accelerometer sensors. For data processing, adequate coding was performed to predict activity according to the pregnant woman’s record. There are features such as chatbot (conversation by messages) and voice assistant. Figure 1 describes the integration of the various components of the system: input (user-computer interface), processing and output/storage of information. Raspbian (Raspberry Pi 3) was used to integrate the cell phone with the other equipment. In the hardware, biosensor kits were purchased for Raspberry Pi or Arduino, which helped to compare them and sensitivity tests between the wristband and sensor kit. The VA was integrated with a device for monitoring pregnant women (ActigraphGT3X) Actigraph (2012). For this integration, it is important for the equipment to contain a communication API, so that the AV has a real-time communication link with other devices to share data. Alternativaly, the data is stored to locally and then retrieved after a certain period of time.

**Figure 1.**
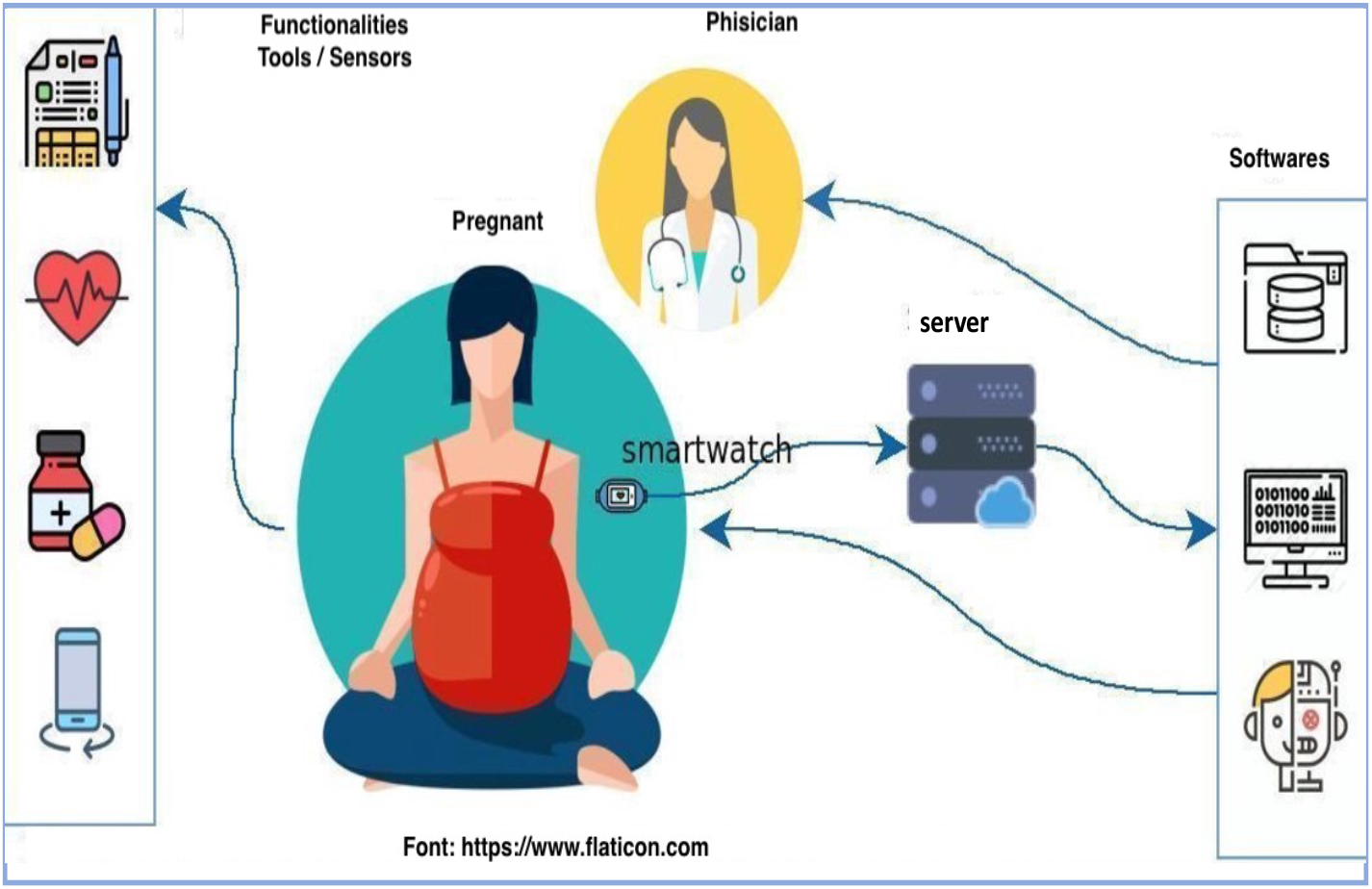
Illustration of Virtual Assistant processing

#### Phase IV: Pilot Study

The pilot study was carried out in two health facilities in the city of Ribeirão Preto, São Paulo State between October and November 2020, which at this time, due to the COVID-19 Pandemic, were carrying out prenatal consultations. Thirty pregnant women who used the Unified Health System-UHS (called by SUS in Brazil) in the city of Ribeirão Preto, Brazil participated in the study. Pregnant women at various different gestational ages was randomly selected. The objective of the pilot study was to evaluate the adequacy of the study design, verify the adaptations made in the PPAQW questionnaire and the functioning of the virtual assistant - EVA version 01-(See details in the EVA construction stage). The prototype of EVA-version 01 counted as a frontend of the MOMeva application developed on an Android that can be installed on a cell phone via Google Play store at: https://play.google.com/store/apps/details?id=app. momeva. The inclusion criteria were pregnant women over 18 years old, low-risk pregnant women; literate; having a cell phone (android) and Internet access. The exclusion criteria were: any morbidity that made it impossible to perform the usual PA on medical recommendation. Pregnant women under 18 years old were not included because it is considered a pregnancy risk due to the biological changes that are still occurring in the woman’s body (Sattler et al., 2018).

#### Data collection

Data collection included two face-to-face interviews with an interval of seven days between them. The interviewer contacted the BHF to obtain information about the hours and days of the appointments. In the first interview, information was collected on socio-demographic conditions, morbidities, lifestyle, weight and height verified in the pregnant woman’s record, and PA all answered in the MOMeva application. The pregnant woman’s record is an essential instrument to collect and transmit data on the health of the pregnant woman and the baby, which aims to ensure communication between the professionals involved and is made available by the Brazilian Department of Health (Dos Santos Lima et al., 2020).

The accelerometers were delivered to the pregnant women packed in plastic bags that had been properly sanitized, along with a leaflet containing information on using the device and a contact phone number to call in case of questions or technical problems about the device. The interviewers used a tablet/cell phone with internet access in case the pregnant woman did not have access to the internet on her cell phone. Eligible pregnant women were instructed to use the accelerometer (Actigraph GT3X) (Actigraph, 2012), for seven days, uninterrupted, on the non-dominant wrist. The study adopted the monitor on the wrist instead of the traditional method where it is located at the waist. This choice was based on scientific evidence which demonstrated that wrist-worn monitors can accurately estimate PA throughout a day routine and increases the rate of use among participants. It is also more comfortable for pregnant women to use them related to hip positioning (Souza et al., 2019; Sasaki et al., 2017). Participants did not need to press any button on the device. The device was configured to record data from PA movements automatically based on when they were delivered to the participants in the HFs. The battery charge level was checked by the technical team before handing over the device.

During the week, the pregnant woman had to use the MOMeva application to record the type of activity she was doing. On the first page of the application there were some pictures of activities that the pregnant woman could be doing at that time, therefore the participants had to click on the picture of when the activity started and then click again when they finished, Figure 2

**Figure 2.**
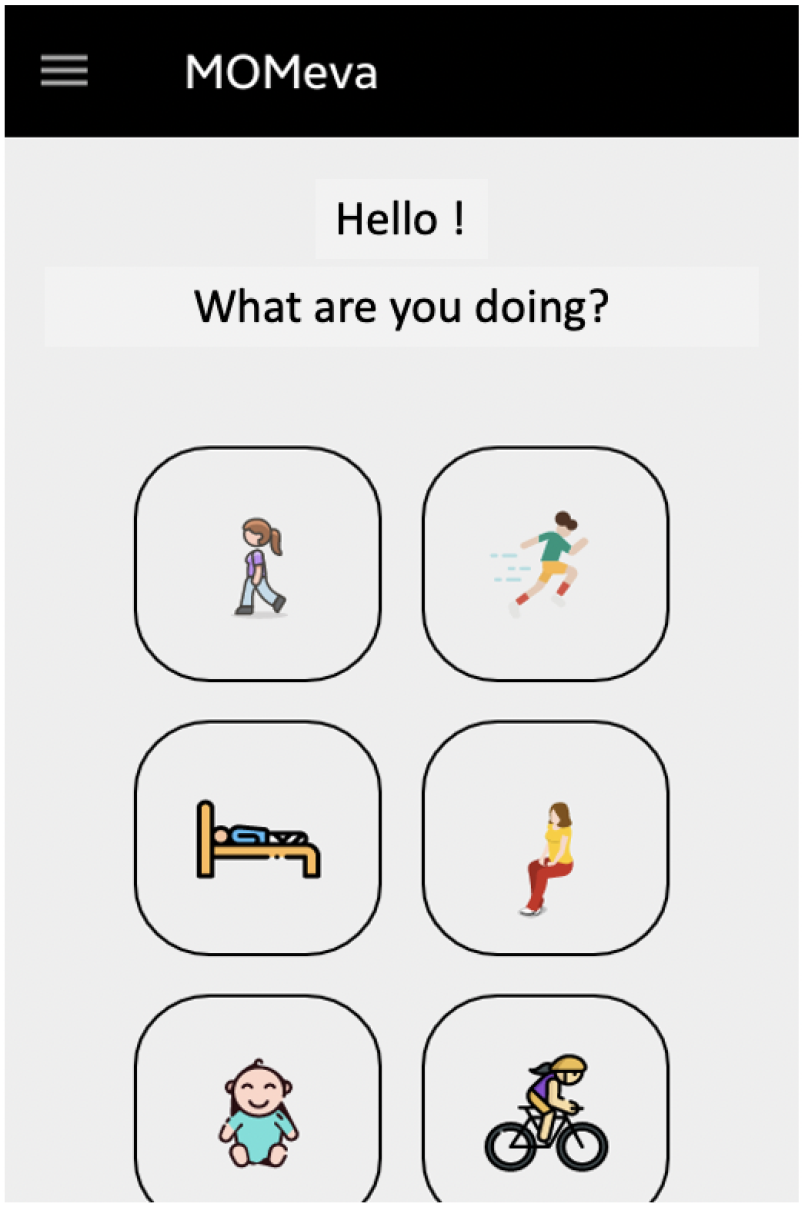
Homepage of App MonEVA

After seven days from the first interview, the participants answered the PA questionnaire again proposed in the VA and the accelerometers were collected by a team member. Their contents were downloaded in the cloud to integrate them with the records made about the PA type in the app.

#### Variables

All research procedures were clarified and the pregnant woman was invited to answer the PA question-naires, screening questionnaire, sociodemographic data, morbidities, lifestyle and nutritional status in the MOMeva application^1^.

The socio-demographic variables verified were: Age in years, self-reported skin color, marital status, education (of the pregnant woman and the head of household), family income and possession of consumer goods, head of household (that considers who contributes with the highest income in the household), consumption of alcoholic beverages, smoking, presence of obstetric history, morbidities, family history of chronic diseases (diabetes, hypertension); Work outside the home (yes or no); Occupation (cleaning assistant, cleaner, secretary or clerk and other activities); social class: For the economic classification, the Brazilian Economic Classification Criteria (Brasil, 2019) in 2019^2^, defining the economic classes of A (highest level) to E (lowest level), which considers the education of the head of household, data on possession of goods by the household and public services (piped water and paved roads); Gestational age (To calculate the gestational age, the date of the Last Menstrual Period (LMP) recorded in the pre-natal details was used); Pre-gestational weight, current weight (kg) and height (cm) were reported by the pregnant woman and confirmed by the pre-natal details. Pre-gestational and current Body Mass Index (BMI): obtained by dividing the pre-gestational and current weight divided by the height squared; pre-gestational BMI adequacy: the Institute of Medicine criteria Pregnancy (2009) were adopted to assess the pre-gestational BMI adequacy; BMI adequacy by week gestational period: the criteria proposed by Atalah Atalah Samur et al. (1997) were used, which considers the gestational week of the pregnant woman.

#### Physical Activity

The PA variable was quantified by the adapted PA questionnaire(B-PPAQ adapted after focus group) contained in MOMeva and also by the accelerometer (ActigraphGT3X). In the questionnaire, the time spent (minutes/day) on each activity was verified over the last gestational trimester. For the time spent on each activity per day per minute, we used the METs that estimate the average weekly energy expenditure, see Tavares et al. (2009). We considered the Total Time spent on activities in a week, multiplied by the MET of the activity, called (TTSP). The MET of each physical activity is related to the oxygen consumption (about *V* 0_2_ by 3.5 mL *kg*^−1^*min*^−1^), which varies according to the intensity of the exercise. The METs of each physical activity can be checked in the Physical Activity Compendium (Ainsworth et al., 2000).

#### Accelerometers

To categorize the PA intensity on the accelerometer, we used the Vector of Magnitude Per Minute (VMPM) proposed by Sasaki JE (2011) and Keadle (2014). Table 1 presents the cutoff points given by the authors.

**Table 1.** PA Intensity (VMPM/day) according to cut-off points by the authors.

| PA Intensity | Sasaki(2011) | Keadle(2014) |
| --- | --- | --- |
| Sedentary | - | 0-199 |
| Light | 0-2690 | 200-2689 |
| Moderate | 2691-6166 | ≥ 2690 |
| Vigorous | 6167-9642 | - |
| Very Vigorous | ≥ 9643 | - |

Accelerometers are motion sensors based on accelerometry that are portable and easy to use. The most used models in research are triaxial, sensitive to movements in the three axes (vertical, anteroposterior and lateral) (Souza et al., 2019). The authors, shown in Table 1 considered three axes, 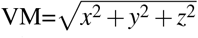. The ActiGraph device model GT3X (Manufacturing Technology, Inc, MTI) (Actigraph, 2012) is similar to a common watch. The device is light and compact, weighing 27 grams and measuring 3.8 cm x 3.7 cm x 1.8 cm and water resistant. The device uses a triaxial accelerometer with a solid sensitive transducer to movements performed between 0.05 to 2.5G in magnitude and epoch can vary from 1 to 360 min. One digital converts the detected movement into an electrical signal in a frequency range between 0.25 up to 2.5 Hz. In this study, the sampling rate at a frequency of 100 Hz was used. Studies recommend that data collection at higher frequencies and reintegrated for lower frequencies in the post-collection period, if necessary. It should be mentioned that data cannot be reintegrated from lower to higher frequencies (Souza et al., 2019; Arvidsson et al., 2019). The result is given in VM related to the intensity and frequency of the movement used. The data generated will be stored and downloaded to a computer via the software using a USB interface cable, provided by the manufacturer.

## STATISTICS ANALYSIS

The questionnaires were collected using the Google Forms application da Silva Mota (2019), whose data are saved in CSV files spreadsheets and later inserted into a database hosted in DBMS MySQL using the R programming language R Core Team (2021).

To verify the reproducibility of the questionnaire, the Student’s t or Wilcoxon test was used to compare two means of TTPS1 and TTPS2. The significance level was *α*= 5% and the statistical analyses were performed using the R Core Team (2021).

The accelerometers were configured and downloaded using the Actilife 6.13.4 software(Actigraph, 2012) and convert the RAW files into CSV files for analysis. For the data analysis generated by the accelerometers, the time that the accelerometer was worn, or wear-time, was at least 5 days a week.

The activity intensity was estimated with the average time per week (hours/week that were later converted into minutes/week) spent on sedentary, light, moderate and intense PA from time series (Epochs) aggregates of 60 seconds.(Arvidsson et al., 2019; Sasaki et al., 2017). The intensity of the activities were calculated by Vector Magnitude per minute in *countsmin*^−1^ (VMPM).

To quantify how many hours the VMPM were within certain cutoff points by sedentary, light, moderate and intense PA. We considered the cutoff given by Keadle (2014) and Sasaki JE (2011), because they considered adults (women and man) and triaxial accelerometers (VM3). To compare the PA estimates from the PPAQ with the PA estimates obtained by the accelerometer, the hours recorded on the forms were considered. The estimated time of PA recorded were [0-30], [31-60], [61-120], [121-180] and [major of 180 min/perday] in seven days (one week) and the average of ranges was considered for each PA category: sedentary, light, moderate and intense. For the VMPM analysis, the cutoffs (Keadle (2014) and Sasaki JE (2011)) and the hours recorded in the accelerometer, were considered. Basically, we counted how many hours were spent on each cut-off point and compared these results with how many hours were spent on each categories sedentary,light, moderate and intense, by METS. These categories were defined by each PA on B-PPAQ Sasaki JE (2011).

## ETHICS AND DISSEMINATION

All available data were analyzed maintaining the identity of the participants confidential, as recommended by Resolution CNS 466/12. This study was approved by the Research Ethics Committee at the School Health Center, Ribeirão Preto Medical School, University of São Paulo (3,681,225) and by the Ribeirão Preto Department of Health. The pregnant women who agreed to participate in the study signed the informed term of consent contained in the MOMeva application.

## RESULTS

### Focus Group

The main findings of the focus group were: 1) Pregnant women reported spending more time performing household and sedentary activities, because they did not feel well due to pregnancy; 2) There is a strong presence of popular beliefs regarding PA in pregnancy, which limits pregnant women doing PA; 3) Among pregnant women who had an occupation outside the home, more than half reported being a cleaner or babysitter.

### Adapting the questionnaire

Based on the focus group observations and after experts discussed the topic, no major adaptations were necessary in the PPAQW, Table 2. Small adjustments were made in the verb tenses (from the infinitive to the third person) at the beginning of the questions. Questions were also included regarding the use of technologies such as using cell phones, tablets, notebooks that were not included in the PAQP and PPAQ. The questionnaire adapted by this study was called the B-PPAQ.

**Table 2.**
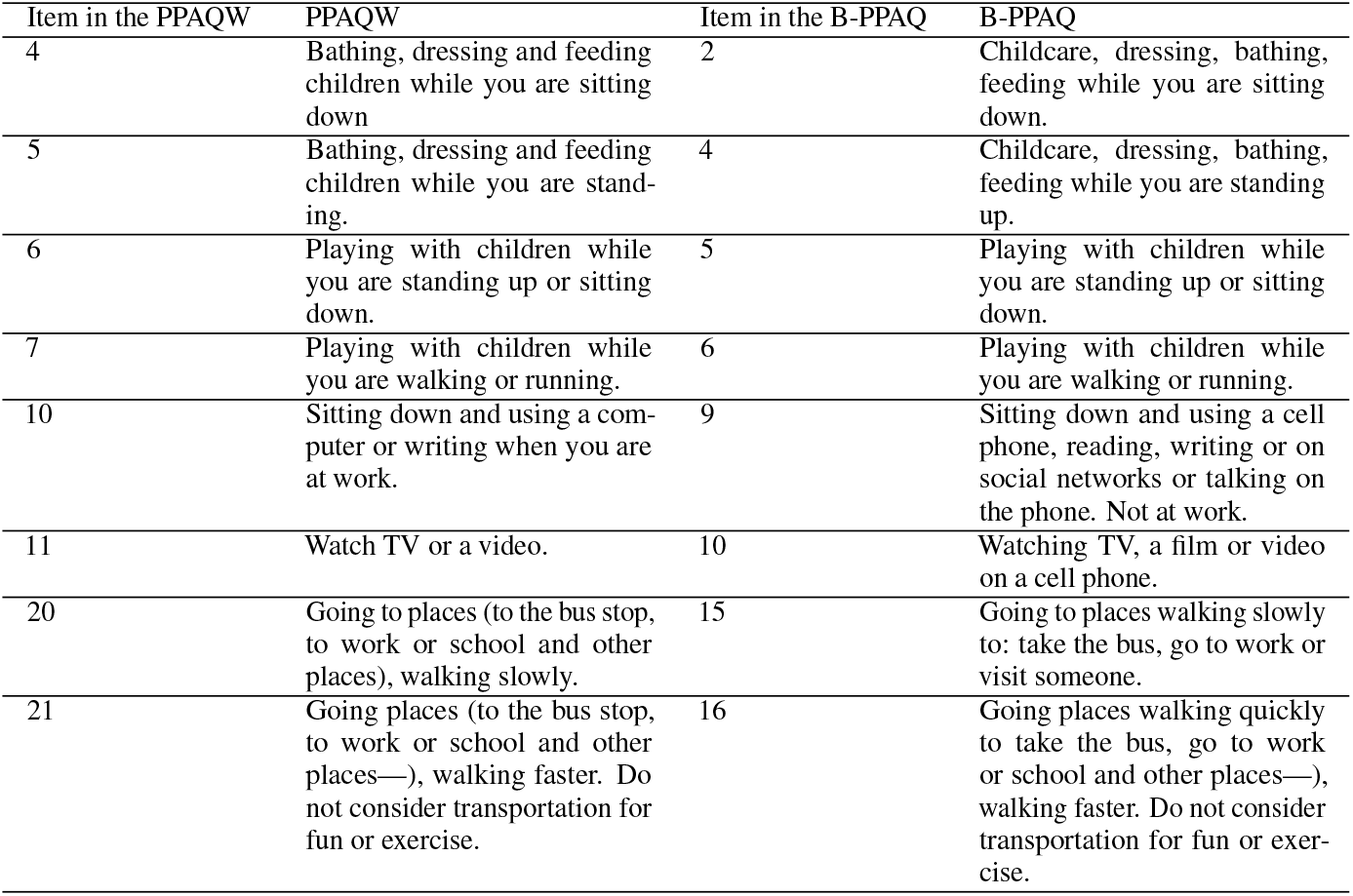
Modification to the PPAQW questionnaire according to the focus group.

| Item in the PPAQW | PPAQW | Item in the B-PPAQ | B-PPAQ |
| --- | --- | --- | --- |
| 4 | Bathing, dressing and feeding children while you are sitting down | 2 | Childcare, dressing, bathing, feeding while you are sitting down. |
| 5 | Bathing, dressing and feeding children while you are standing. | 4 | Childcare, dressing, bathing, feeding while you are standing up. |
| 6 | Playing with children while you are standing up or sitting down. | 5 | Playing with children while you are standing up or sitting down. |
| 7 | Playing with children while you are walking or running. | 6 | Playing with children while you are walking or running. |
| 10 | Sitting down and using a computer or writing when you are at work. | 9 | Sitting down and using a cell phone, reading, writing or on social networks or talking on the phone. Not at work. |
| 11 | Watch TV or a video. | 10 | Watching TV, a film or video on a cell phone. |
| 20 | Going to places (to the bus stop, to work or school and other places), walking slowly. | 15 | Going to places walking slowly to: take the bus, go to work or visit someone. |
| 21 | Going places (to the bus stop, to work or school and other places—), walking faster. Do not consider transportation for fun or exercise. | 16 | Going places walking quickly to take the bus, go to work or school and other places—), walking faster. Do not consider transportation for fun or exercise. |

The most relevant changes made to the questionnaire adapted were in relation to the household and child care. The aforementioned study found an overestimation of energy expenditure by these pregnant women concerning the occupational domain. As it was observed during the focus group that the occupations most reported among pregnant women who worked outside the home were household, cleaning or babysitting, it was decided to adapt the PA questionnaire for pregnant women with these occupations. Thus, when these women responded that they had these occupations, they were automatically asked to answer the PA questionnaire in the occupational domain, which in this case was a copy of the household domain. When they finished responding, they were redirected back to complete the questionnaire Silva et al. (2021).

### Pilot Study

Thirty pregnant women participated, 26 women were considered elegible, Table 3. The characteristics were most pregnant women were aged between 18 and 30 years and had adequate weight. No pregnant woman had, or previouslyhad, any infection and/or gestational diabetes.

**Table 3.**
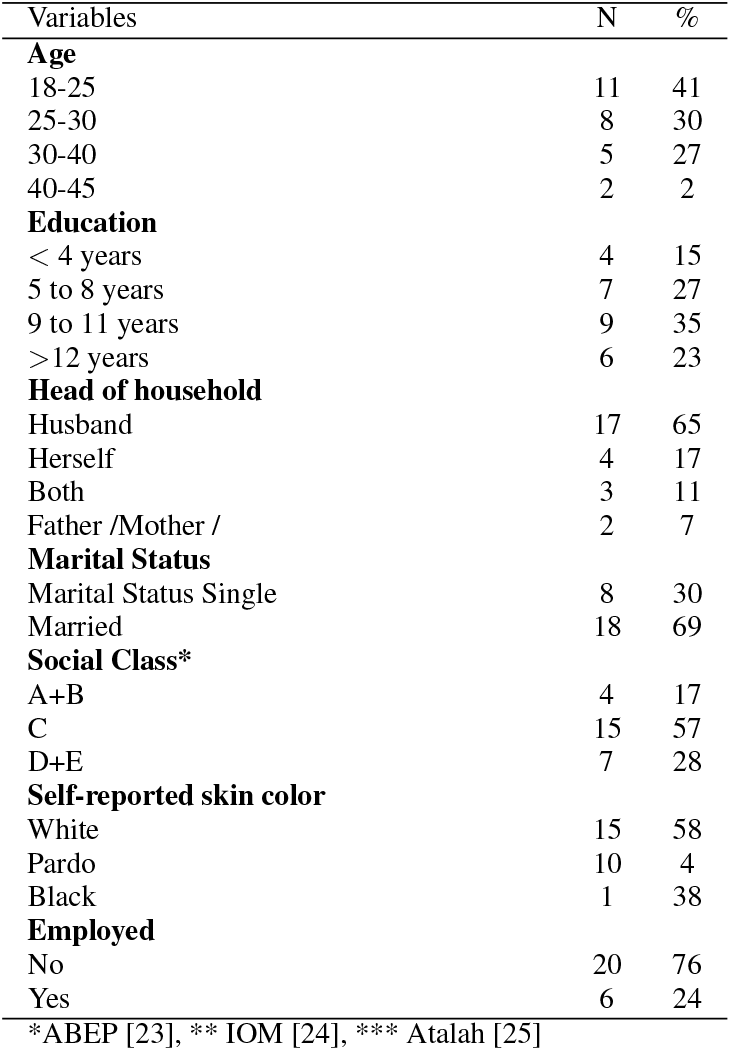
Sociodemographic characteristics of pregnant women.

**Table 4.**
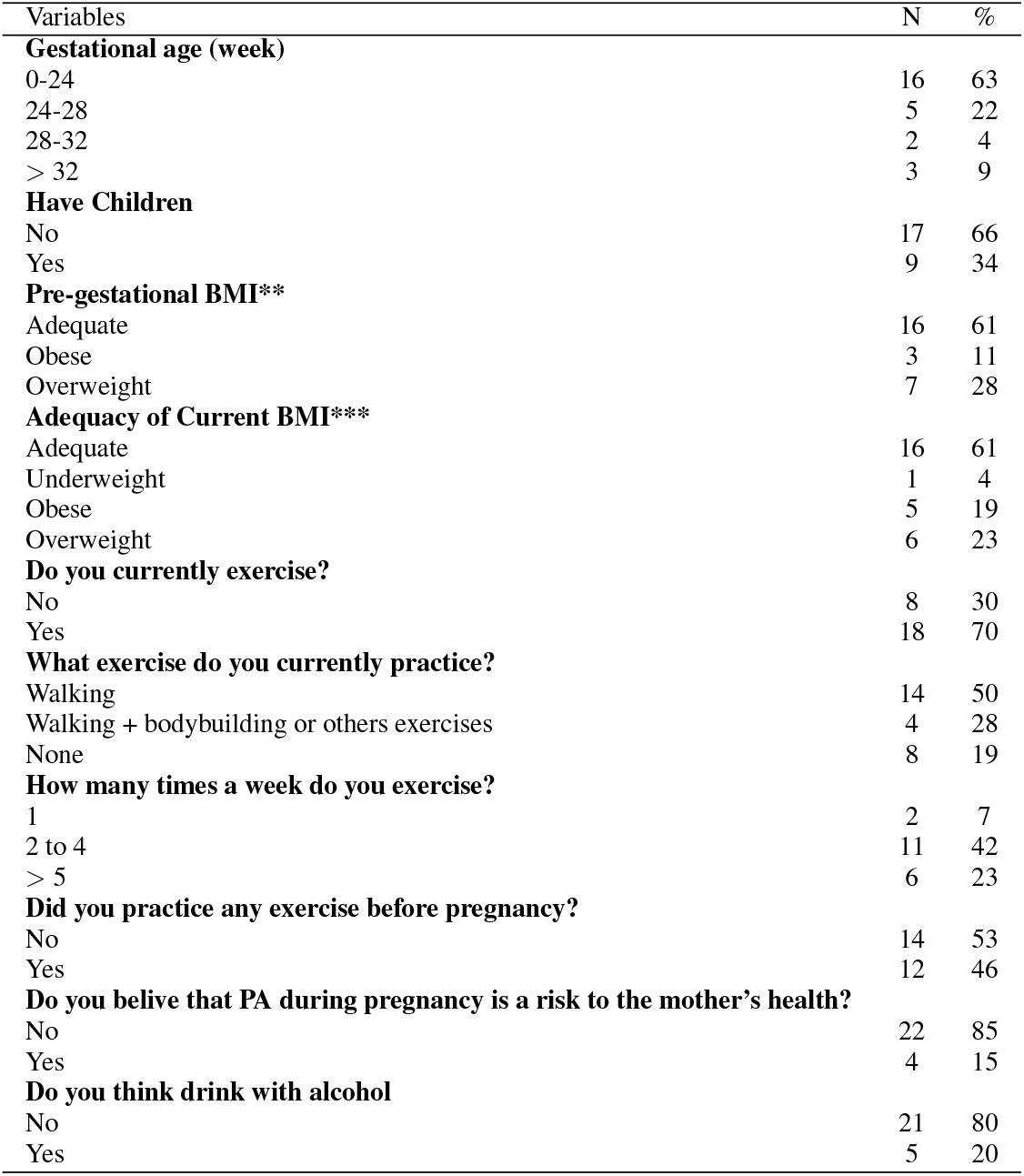
Nutritional and obstetric status and lifestyle of pregnant women.

Concerning time of energy expenditure (TTSP) total in the PA domains, Table 5, shows that the TTSP is greater for household domain.

**Table 5.** Time of the energy expenditure of pregnant women (TTSP) in hours per week for two times self-administered Pregnancy Physical Activity Questionnaires Women adapted (B-PPAQ) by domains and intensity levels and among 26 pregnant subjects. Validation phase, Ribeirão Preto,2022

| Types of PA | Total(TTSP<sub>1</sub>) | Total(TTSP<sub>2</sub>) | Mean<sub>1</sub> | Mean<sub>2</sub> | MED<sub>1</sub> | MED<sub>2</sub> | SD<sub>1</sub> | SD<sub>2</sub> | p-value |
| --- | --- | --- | --- | --- | --- | --- | --- | --- | --- |
| **By Domain** |  |  |  |  |  |  |  |  |  |
| Household (n=26) | 2764.0 | 2972.5 | 106.3 | 114.3 | 81.0 | 99.4 | 6.9 | 88.6 | 0.4195 |
| Ocupational (n=6) | - | 327.9 | - | 65.5 | - | 41.3 | - | 36.5 | - |
| Leisure(n1=19,n2=18) | 686.9 | 594.1 | 36.2 | 22.9 | 18.4 | 15.8 | 63.7 | 25.9 | 0.4410 |
| **By Intensity** |  |  |  |  |  |  |  |  |  |
| Sedentary (n=26) | 1032.9 | 648.9 | 39.7 | 24.9 | 41.3 | 24.1 | 14.2 | 14.4 | 0.0012 |
| Light (n=26) | 2183.3 | 1976.5 | 84.00 | 81.8 | 68.0 | 69.1 | 45.5 | 67.5 | 0.0637 |
| Moderate (n<sub>1</sub>=26 ; n<sub>2</sub>=25) | 1491.9 | 1907.3 | 57.4 | 76.0 | 34.3 | 58.6 | 69.7 | 61.9 | 0.0696 |
| Vigorous(n<sub>1</sub> = 5; n<sub>2</sub> = 11) | 203.0 | 205.7 | 40.6 | 21.31 | 12.2 | 10.5 | 54.3 | 15.0 | 0.5862 |
| PA Total by intensity | 4911.1 | 4415.2 | 174.1 | 178.0 | 131.2 | 151.1 | 122.3 | 121.2 | 0.9403 |

MED1 e SD1: Median and Standard Deviation at the first interview and MED2 and SD2 after 7 days. pvalor < 0.05 to Wilcoxon test of two times. The categories Sedentary (<1.5 METs), Light (1.5 - 3.0 METs),Moderate (3.0 - 6.0 METs) and Vigorous (>6.0 METs)

The locomotion and occupational domains had the lowest average and activities recorded by pregnant women. The questionnaire was applied twice, showing its repeatability for the domains and intensities, only sedentary activities had significant differences between the means of the TTSP. Regarding activity intensities, pregnant women had higher TTSP in light and moderate activities, respectively.

The Table 6 show the hours week recorder by accelerometer by intensity levels to 17 pregnant, nine pregnant did not record into the actigraph. The hours were obtained to considering the cut-off of the authors Keadle (2014) and Freedson (1998). To compare the B-PPAQ and the accelerometer register we considered the hours per week for both. For B-PPAQ, the mean of sedentary was 26.04 hours, light 21.51 moderate 23.47 and intense 1.54 hours per week. The average of the hours obtained by the PPAQW is lower than that obtained by the accelerometer, indicating that pregnant woman underestimates the activities performed.

**Table 6.** Weekly hours per accelerometer by intensity levels, for the 17 pregnant women in the pilot study. Ribeirão Preto,2022.

| Pregnant | Total of hours | Keadle |  |  | Freedson |  |  |  |
| --- | --- | --- | --- | --- | --- | --- | --- | --- |
|  |  | Sedentary | Light | Moderate | Light | Moderate | Intense | Very intense |
| 1 | 93.08 | 62.2 | 17.73 | 13.13 | 79.95 | 10.2 | 1.85 | 1.07 |
| 2 | 139.02 | 69.73 | 22.65 | 46.62 | 92.4 | 28.55 | 15.67 | 2.37 |
| 3 | 150.02 | 74.82 | 38.53 | 36.65 | 113.37 | 24.55 | 10.27 | 1.83 |
| 4 | 139.02 | 69.52 | 33.35 | 36.13 | 102.88 | 22.93 | 11.12 | 2.02 |
| 5 | 139.02 | 64.47 | 42.12 | 32.38 | 106.63 | 24.55 | 6.73 | 1.1 |
| 6 | 139.02 | 67.1 | 29.22 | 42.68 | 96.33 | 29.9 | 11.62 | 1.15 |
| 7 | 139.02 | 71.75 | 43.82 | 23.45 | 115.57 | 17.85 | 4.23 | 1.37 |
| 8 | 146.02 | 80.22 | 33.92 | 31.88 | 114.13 | 20.92 | 9.62 | 1.35 |
| 9 | 139.02 | 64.93 | 29.68 | 44.33 | 94.68 | 29.43 | 12.07 | 2.82 |
| 10 | 139.02 | 104.92 | 21.3 | 12.78 | 126.23 | 9.35 | 3.02 | 0.4 |
| 11 | 139.02 | 70.95 | 34.7 | 33.35 | 105.67 | 25.8 | 7.08 | 0.47 |
| 12 | 148.02 | 125.35 | 11.62 | 11.05 | 136.97 | 7.88 | 2.88 | 0.28 |
| 13 | 146.02 | 59.85 | 37.92 | 48.23 | 97.78 | 25.2 | 15.25 | 7.77 |
| 14 | 146.02 | 75.68 | 38.08 | 32.22 | 113.8 | 13.68 | 11.47 | 7.07 |
| 15 | 155.02 | 99.17 | 30.87 | 24.95 | 130.07 | 16.05 | 7.02 | 1.88 |
| 16 | 148.02 | 126.4 | 9.38 | 12.23 | 135.78 | 8.6 | 3.28 | 0.33 |
| 17 | 146.02 | 74.03 | 46.02 | 25.92 | 120.1 | 18.63 | 6.37 | 0.88 |

Keadle, Shiroma,Freedson, et al(2014): Sedentary (VM cpm <199), Light (VM cpm 200-2689), Moderate (VM cpm ≥2690) and Sasaki, JE; John, D; Freedson, PS.(2011): Light (VM cpm <2689), Moderate (VM cpm 2690 – 6166), Vigorous (VM cpm 6167 – 9642) and Very Vigorous (VM cpm ≥ 9643)

**Table 7.** Weekly hours per accelerometer by intensity levels, for the 17 pregnant women in the pilot study, record on B-PPAQ questionaire and by the categories. Ribeirão Preto,2022.

| Pregnant | by B-PPAQ |  |  |  | by acelerometer(Total) | by B-PPAQ (Total) |
| --- | --- | --- | --- | --- | --- | --- |
|  | Sedentary | Ligh | Moderate | Intense |  |  |
| 1 | 22.75 | 42.00 | 35.00 | 0 | 93.08 | 99.75 |
| 2 | 22.75 | 3.50 | 21.00 | 0 | 139.02 | 47.25 |
| 3 | 35.00 | 5.25 | 3.50 | 0 | 150.02 | 43.75 |
| 4 | 19.25 | 14.00 | 22.75 | 1.75 | 139.02 | 57.75 |
| 5 | 35.00 | 24.50 | 10.50 | 1.75 | 139.02 | 71.75 |
| 6 | 22.75 | 15.75 | 19.25 | 0 | 139.02 | 57.75 |
| 7 | 42.00 | 3.50 | 10.50 | 0 | 139.02 | 56.00 |
| 8 | 35.00 | 8.75 | 10.50 | 0 | 146.02 | 54.25 |
| 9 | 10.50 | 31.50 | 12.25 | 0 | 139.02 | 54.25 |
| 10 | 10.50 | 8.75 | 3.50 | 0 | 139.02 | 22.75 |
| 11 | 22.75 | 19.25 | 49.00 | 0 | 139.02 | 91.00 |
| 12 | 26.25 | 19.25 | 29.75 | 0 | 148.02 | 75.25 |
| 13 | 35.00 | 33.25 | 82.25 | 22.75 | 146.02 | 173.25 |
| 14 | 22.75 | 14.00 | 7.00 | 0 | 146.02 | 43.75 |
| 15 | 19.25 | 19.25 | 15.75 | 0 | 155.02 | 54.25 |
| 16 | 19.25 | 94.50 | 63.00 | 0 | 148.02 | 176.75 |
| 17 | 42.00 | 8.75 | 3.50 | 0 | 146.02 | 54.25 |
| Mean | 26.04 | 21.51 | 23.47 | 1.54 | 140.61 | 72.57 |
| SDesv | 9.70 | 21.73 | 22.39 | 5.50 | 13.23 | 42.51 |

Sedentary (<1.5 METs), Light (1.5 - 3.0 METs),Moderate (3.0 - 6.0 METs) and Intense (>6.0 METs)

**Table 8.** Percentage (10%,50% and 90%) of total of the VM3 (*countsmin*^−1^) of 17 pregnant women by accelerometer. Ribeirão Preto,2022.

| Pregnant | P10 | P50 | P90 | Mean |
| --- | --- | --- | --- | --- |
| 1 | 179.3 | 1808.2 | 5665.4 | 2605.8 |
| 2 | 356.9 | 4039.3 | 7917.0 | 4257.3 |
| 3 | 137.3 | 1994.5 | 6934.9 | 2905.5 |
| 4 | 188.0 | 2411.1 | 7288.0 | 3258.0 |
| 5 | 209.1 | 1954.0 | 6077.2 | 2663.2 |
| 6 | 335.1 | 3070.5 | 6953.3 | 3447.6 |
| 7 | 149.7 | 1501.0 | 5532.4 | 2323.7 |
| 8 | 140.2 | 2050.1 | 6965.6 | 2918.1 |
| 9 | 231.4 | 3137.2 | 7668.8 | 3639.7 |
| 10 | 140.8 | 1686.7 | 5869.7 | 2403.6 |
| 11 | 198.0 | 2272.1 | 6165.6 | 2808.3 |
| 12 | 59.0 | 1972.0 | 6490.6 | 2682.2 |
| 13 | 251.0 | 2787.4 | 9019.7 | 3881.2 |
| 14 | 94.0 | 1644.8 | 9176.4 | 3288.2 |
| 15 | 100.1 | 1875.3 | 7052.2 | 2812.9 |
| 16 | 745.2 | 3082.5 | 7062.5 | 3543.8 |
| 17 | 93.1 | 1432.0 | 5825.0 | 2263.6 |
| Mean | 212.2 | 2277.6 | 6921.4 | 3041.3 |
| SD | 159.7 | 714.9 | 1075.2 | 569.9 |

( $\leq 211$ ) Sedentary (212 – 2278) Light (2279 – 6921) Moderate(> 6922) Intense

## DISCUSSION

The objective of this work is to present the results of the adaptation of the PPAQW questionnaire and to present the results of the pilot study for the construction of the Virtual Assistant (EVA) in Portuguese, providing an example of a health management resource for monitoring the physical activities of pregnant women assisted in the Unified Health System. The main findings related to the adaptation of the questionnaire were the inclusion of questions related to the use of cell phones, tablets and notebooks and the adaptation of the occupational domain in pregnant women whose occupation is domestic work (cleaning lady, nanny). And the determination of cut-off points to classify PA intensities in pregnant women using the accelerometer(GT3X activity monitors), were also observed. The choice of the PPAQW, to adapt it for this work, was based on a previous literature review in which the PPAQW showed good cross-cultural validity based on the criteria selected by the study. Issues related to the use of cell phones, tablets and notebooks were not found in the PPAQW, as well as in the initial version of the PPAQ. However, recently the PPAQ was updated and these questions related to contemporary sedentary behavior were included Chasan-Taber et al. (2023).Sedentary behavior involves activities performed when awake sitting, reclining or lying down and expending little energy. Scientific evidence shows that long periods of sedentary behavior are related to a greater risk of complications during pregnancy and childbirth. The adaptation made to our proposal, regarding the occupational domain of maids, cleaners and nannies, was based on previous research, in which the TTSP of pregnant women with a domestic occupation was overestimated (Da Silva et al, 2021) and, in the focus group, where pregnant women who did not work declared that they had this type of occupation previously. However, it was not possible to verify whether this adaptation changes the classification of pregnant women in terms of intensity due to the small number of the pilot sample. However, we consider it important that this hypothesis is evaluated when developing this project. No other study with pregnant women mentioned this issue. A possible reason for this is that domestic occupation is higher in Brazil than in other countries. The results observed in the pilot study, unlike other research Bernardo (2019); Silva et al. (2021); Papazian et al. (2020); Silva et al. (2015); Siyad et al. (2022) and questioning the hypothesis that pregnant women are a sedentary population, found that the majority of pregnant women in this study performed light activities followed by moderate and sedentary, respectively. In relation to the domains, the domestic domain had the highest TTSP, corroborated by other studies Bernardo (2019); **?**); Papazian et al. (2020); Silva et al. (2015); Siyad et al. (2022). While the occupational domain had the lowest energy expenditure due to the lower number of pregnant women (76%), (transport domain) In this study, it was found that only sedentary activities showed significant differences between the questionnaires, which showed good repeatability in general. However, this fact shows that the difference of one week can limit sedentary observation. Comparing the PA intensity results from the questionnaire with the results obtained by the accelerometer, it was found that the average total PAs verified in the questionnaires were lower than those observed by the accelerometers, indicating that women underestimate the activities. Although the sample number is small, these results corroborate other studies such as Chasan-Taber et al. (2023) and Santos et al. (2023) (in which some activities were underestimated, but others were overestimated. Regarding cutoff points, there is a discussion in the literature of which cutoff points provide the best comparison for this population, as the cutoff points found in studies have not been validated for pregnant women Sattler et al. (2018). In this study, the cutoff points of Keadle (2014) and Sasaki JE (2011) were used, because they considered adults (women an male) and triaxial accelerometer (VM3). According to these authors, pregnant women are classified as sedentary. Despite the limitation regarding the sample size, this study managed to establish new cut-off points for pregnant women for the intensity of activities in VM3 of the accelerometers. There are limitations regarding potential confounding factors, such as the change in participants’ PA when using the accelerometer. Another limitation may be the use of the application and access to the Internet. It is known that in underdeveloped countries’ access to the internet is more limited and the educational level of pregnant women is lower than in developed countries and, therefore, there is the possibility of encountering difficulties in using applications. However, we believe that the development of a VA in Portuguese can bring many benefits to health management in the country, along with PA monitoring, which can be carried out by both the doctor and the pregnant woman herself. Considering the advancement of technology, smartphones have become indispensable for most people, and consequently the applications they use aim to make people’s daily lives easier, both in simple and more complex tasks and can help with health care, as well as in disease prevention and control Carvalho et al. (2019).

## RECOMMENDATION

We recommend further studies so that it is possible to investigate the relevance of adaptations in the occupational domain of pregnant women who work as domestic workers (houseworkers and/or nannies) and with the initial cut-off points established by this study.

## CONCLUSION

This study made important contributions to evaluating PA in pregnant women. The proposal and studies for the construction of the AV-EVA, the inclusion of a specific occupational domain for pregnant women with domestic occupations and the new cutoff points for PA intensity measurements obtained via accelerometers. Despite the limitations, this study addressed issues to be explored in future research, facilitating comparability between studies from different countries. This project will continue for a larger number of pregnant women, 200 pregnant women, as planned to complete the Virtual Assistant.

## Data Availability

All data generated in this study are available from the authors upon reasonable request.

https://eva.fmrp.usp.br/

## ACKNOWLEDGMENTS

We would like to thank FAPESP(Number 2019/03984-8 FAEPA (Fund. de Apoio ao Ensino, Pesquisa e Assistencia do HCFMRP-USP), CAPES the Ribeirão Preto Health Department, Prof. Patricia, as well as the pregnant women who took part in this phase of the study.

## Footnotes

1 https://eva.fmrp.usp.br/momeva-aplicativo-do-projeto-e-desenvolvido/

2 https://www.abep.org/criterio-brasil

